# Weekly changes in ventilation response for photon and proton lung cancer patients during radiotherapy

**DOI:** 10.1101/2025.08.28.25334578

**Authors:** Rebecca Lim, Caleb O’Connor, Joshua Pan, Tien T Tang, Austin H Castelo, Yulun He, Uwe Titt, Radhe Mohan, Zhongxing Liao, Kristy K Brock

## Abstract

**Purpose:** Conformal dose distributions in proton radiotherapy promise to reduce normal tissue toxicity such as radiation-induced pneumonitis, but this has not been fully realized in clinical trials. To further investigate dose and toxicity, we employ voxel-based normal tissue evaluation techniques such as ventilation maps throughout treatment. We hypothesize that ventilation change after 1 week of treatment (WK1) predicts for ventilation change at the end of treatment (EOT).

**Methods:** For 48 photon and 23 proton lung cancer patients, 4DCT-based ventilation maps were generated using stress-based methods at planning, WK1, and EOT. Voxel-wise ventilation change from planning to WK1 and EOT was calculated and binned by planned dose, and median ventilation change at WK1 and EOT was calculated across all patients in each dose bin. Patients were stratified into 6 groups based on modality and increased, decreased, or stable ventilation at WK1. Mann-Whitney U tests were performed to determine if median ventilation change at WK1 and EOT in each dose bin was significantly different from zero. Univariate analysis was performed to correlate ventilation change at EOT with change at WK1 and other clinical factors. A linear regression model was developed to predict ventilation at EOT using a variety of input features including ventilation at planning, ventilation at WK1, tumor response information, and tumor location. Accuracy of the model was assessed through R^2^.

**Results:** For patients that decreased in ventilation at WK1, 90% of photon patients and 92% of proton patients were stratified similarly at EOT. Patients that were stratified as increased ventilation at WK1 were stratified similarly (72% for photon, 80% for proton) at EOT. These patients were more likely to develop Grade 2+ pneumonitis though the difference was not significant when computing a Fisher’s exact test. Univariate analysis indicated that only ventilation change at WK1 was correlated with ventilation change at EOT. The linear regression model achieved R^2^ of 0.65.

**Conclusion:** Ventilation changes at EOT can be predicted using ventilation information from planning and WK1. Patients that increased in ventilation at WK1 were more likely to develop pneumonitis. Further work is needed to characterize the relationship between ventilation change with pneumonitis development.

## Introduction

Lung cancer is the leading cause of cancer death in the United States, with non-small-cell lung cancer (NSCLC) accounting for 85% of these [1-3]. External beam radiation therapy (RT) with concurrent chemotherapy is a common treatment for NSCLC. RT is non-invasive and allows for local tumor control, but the therapeutic potential is typically limited due to the radiosensitivity of the lungs [3, 4]. Ventilation imaging may be performed prior to RT to provide insight into regional lung function. Examples of clinical ventilation imaging modalities are single photon emission computed tomography (SPECT) and positron emission tomography (PET). However, these modalities are unideal for monitoring lung function throughout treatment because they are not standardly acquired during RT, are not imaging modalities typically present in radiation oncology departments, and suffer from issues such as aerosol clumping in airways [5]. Consequently, changes in local lung function is not easily assessed nor quantified throughout the course of treatment.

CT-derived ventilation imaging (CTVI) has emerged as an alternative method for measuring regional lung function by calculating ventilation from four-dimensional CT (4DCTs) [6]. Since 4DCTs are acquired as standard-of-care during radiotherapy for treatment planning and tumor motion assessment and CT scanners are present in most radiation oncology departments, CTVI is an efficient and cost-effective alternative to measuring ventilation compared to SPECT and PET. This method of ventilation imaging also offers higher spatial resolution and presents opportunities for longitudinal studies since 4DCTs are easier to acquire. A variety of techniques have been developed to calculate ventilation from 4DCTs, most commonly using a density-based or an intensity-based algorithm [7-12]. These techniques have been validated in multiple studies against clinically accepted measures of pulmonary function, ranging from SPECT and PET [13-16] to pulmonary function tests (PFTs) [16] to xenon-CT [7].

Recently, CTVI has been implemented in clinical trials that compare functional lung avoidance (FLA) RT with conventional RT [17-21]. FLA seeks to identify high functioning regions of the lung, defined by high levels of ventilation, and devise a treatment plan that spares these regions. The hypothesis of FLA is that reducing dose to high functioning regions of the lung will result in decreased toxicity, leading to better outcomes and higher patient quality of life after treatment. Previous trials have successfully reduced toxicity using SPECT and PET [22, 23], and recent studies have achieved similar results with CTVI [24]. Separate studies have also looked at toxicity modeling with CTVI to predict radiation-induced pneumonitis, a common limiting toxicity associated with lung RT, based on ventilation and have demonstrated the potential of pre-RT ventilation to predict for toxicity [25-30]. However, these trials have only evaluated pre-RT ventilation and there is potential in characterizing and preserving ventilation in the lung to positively impact patient quality of life.

Existing studies on the effects of radiation on lung ventilation have largely focused on changes 3 months after RT [31-34], and only a few have looked at changes over treatment [32, 35-37]. In these studies, no clear relationship between dose and ventilation change during treatment has been found [36], though it has been shown that changes throughout treatment are correlated with tumor shrinkage [36, 37]. However, these studies were limited by patient cohort sizes of less than 20 patients, and to our knowledge, none have looked at predicting ventilation changes at the end of treatment yet.

In this study, we calculated 4DCT-derived ventilation from three different timepoints: planning, 1 week into treatment (WK1), and end of treatment (EOT). We evaluated patients treated with both photon and proton RT and compared the ventilation changes over the course of treatment. We hypothesize that changes in ventilation at WK1 predict for ventilation changes at EOT.

## Methods

### Patient cohort

Patients were selected retrospectively from a previous clinical trial at our institution which treated 149 locally advanced NSCLC patients with either intensity modulated radiation therapy (IMRT) or passive scattering proton therapy (PSPT) [38]. All patients received concurrent chemoradiation and were treated to 74 Gy in 37 fractions or 66 Gy in 33 fractions. After treatment, patients received follow-up CT scans to aid in diagnosis of pneumonitis, which was scored using the Common Terminology Criteria for Adverse Events (CTCAE), version 3.0. As part of the clinical trial protocol, all patients received weekly 4DCT scans throughout treatment. Exclusion criteria included missing 4DCT images, missing dose plans, and incomplete treatment. Patients were excluded if they did not have a 4DCT taken within the last 5 fractions of treatment. To ensure similar dose effects between the two fractionation schemes, patients were also excluded if the dose delivered by the last 4DCT was less than 64 Gy. A flowchart demonstrating patient exclusion criteria is shown in Figure 1. After applying all exclusion criteria, 48 photon and 23 proton patients were identified for this study. Patient demographics and treatment details for the final cohort are shown in Table 1.

**Table 1.**
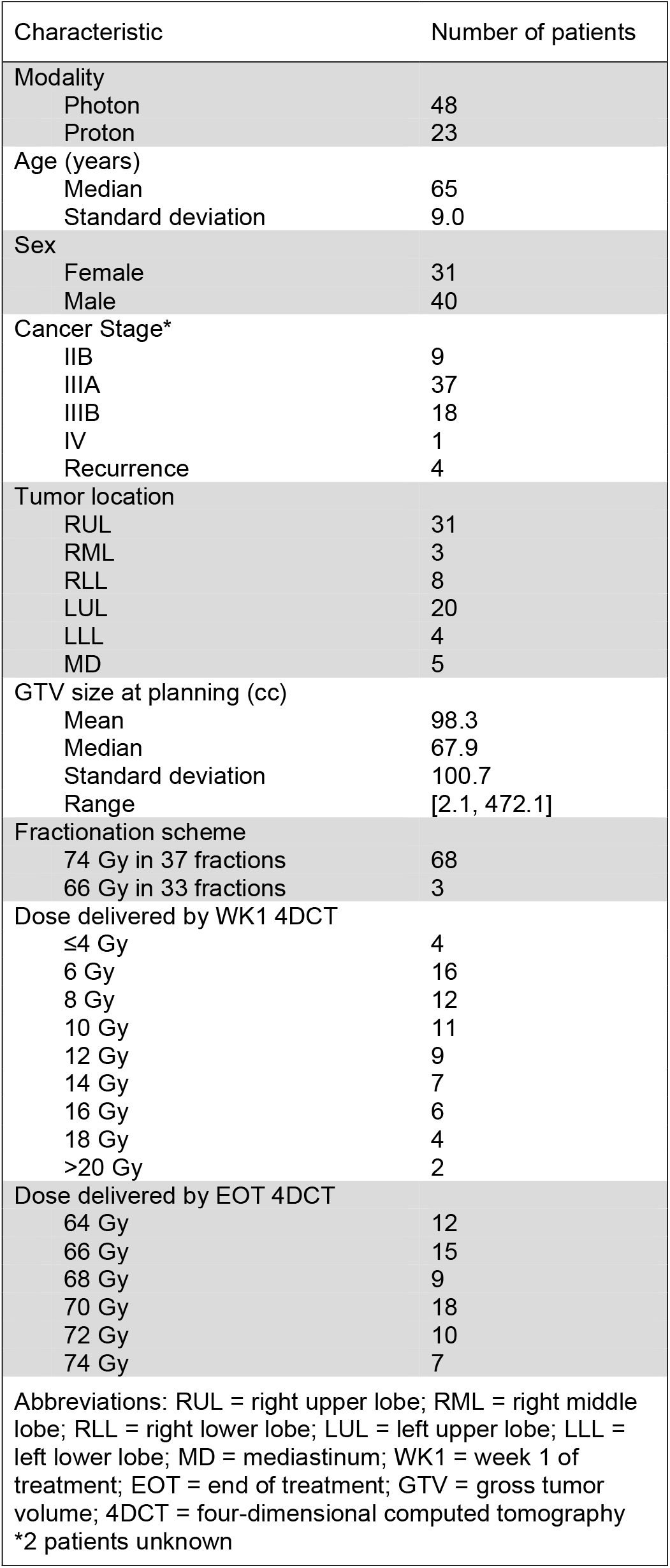
Patient demographics and treatment details.

**Figure 1.**
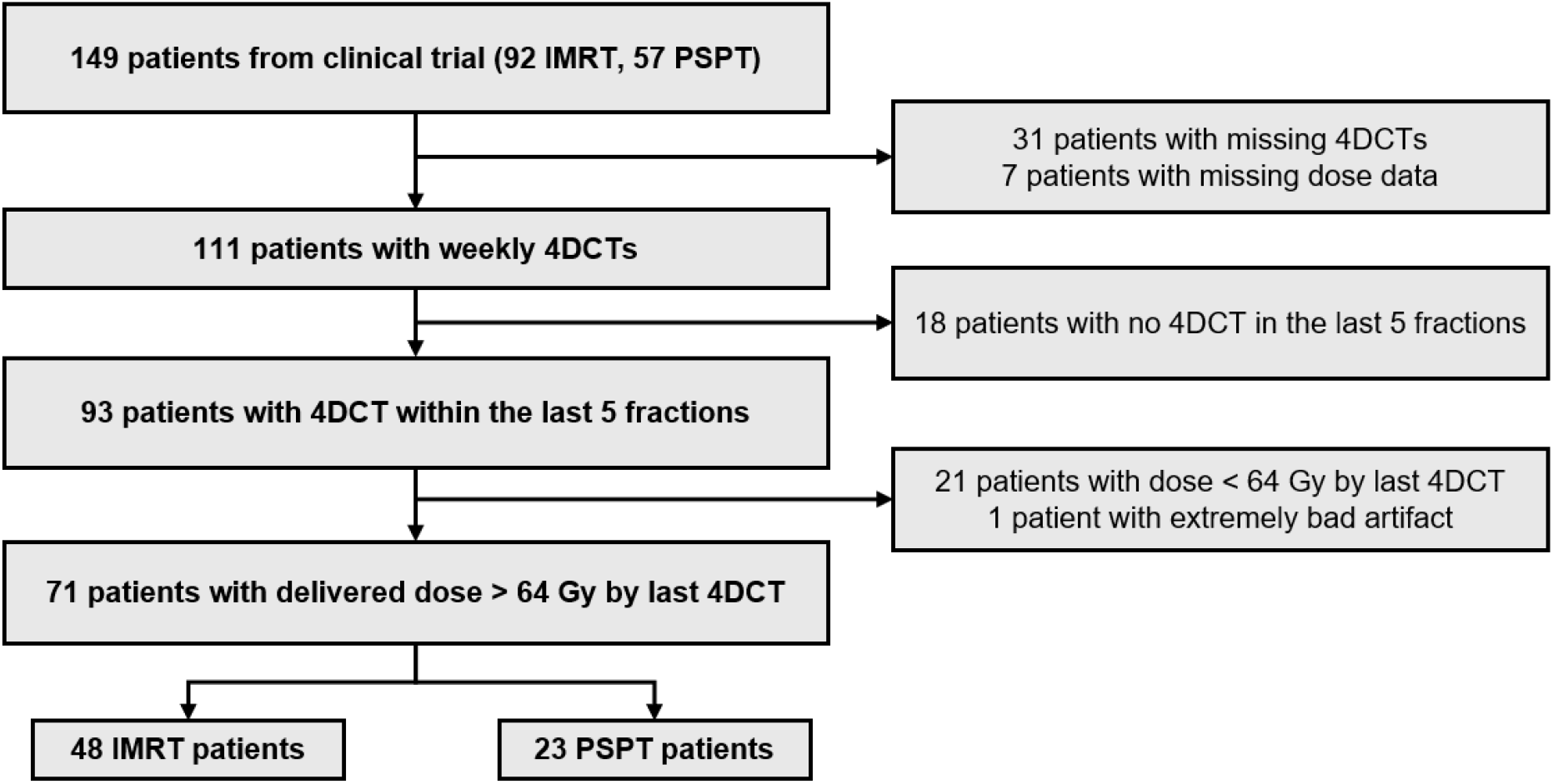
Patient inclusion criteria for this study. IMRT = intensity modulated radiation therapy, PSPT = passive scattering proton therapy, 4DCT = four-dimensional computed tomography

### 4DCT ventilation derivation

4DCTs were acquired weekly for all patients, but this study evaluated only three sets of 4DCTs: at planning, during the first week of treatment, and at the last week of treatment. 4DCTs for each patient were acquired in cine mode with 2.5mm slice thickness on a helical CT scanner (Discovery PET/CT; GE Healthcare, Waukesha, WI).

Maximum principal stress in the lung was calculated as a surrogate for ventilation as previously described in Cazoulat et al. [39]. This algorithm demonstrated superior performance to conventional density-based and intensity-based methods in the multi-institutional VAMPIRE challenge [13] and in an independent validation study [39]. We will briefly summarize the process here.

Deformable image registration (DIR) was performed between the inhale (T0) and exhale (T5) phase images of a 4DCT using a hybrid algorithm that employs both biomechanical and intensity information. This method has been shown to provide superior alignment of lung vessels compared to other algorithms [40, 41]. To compute the deformation, tetrahedral meshes were generated for each lung and the body based on the contours of the reference T5 image. MORFEUS biomechanical DIR [42] was performed to define boundary conditions on the surface of the lungs. Vessels were automatically segmented within the lungs and added to the finite element model as additional boundary conditions. The final deformation was calculated using a finite element analysis solver [43].

To calculate stress, values for Young’s modulus were assigned heterogeneously throughout the lung. Each tetrahedral element of the lung was assigned a value for Young’s modulus based on the HU value of the corresponding voxel in the reference T5 image. Since CT scan resolution was finer than the tetrahedral mesh resolution (5mm), the T5 image was smoothed with a Gaussian filter (σ=6mm) prior to assigning values of Young’s modulus. Finite element analysis was performed to calculate stress at the center of each tetrahedron, and the distribution was then resampled onto the grid of the reference T5 image to produce a ventilation map.

Ventilation maps were computed on 3 sets of 4DCTs for each patient: planning, WK1, and EOT. To align ventilation maps over time, interfraction DIRs were performed with the same deformation algorithm described above. DIR was performed to align the T5 scan of WK1 to the T5 scan of the planning week, and the same was done to align the EOT T5 to the planning week T5. Maps were resampled to 3x3x3 voxel sizes to account for uncertainty in DIR [44]. The deformation vector fields (DVFs) were extracted from the DIRs and applied to the ventilation maps to align them on a voxel-wise basis. A summary of this entire workflow is shown in Figure 2.

**Figure 2.**
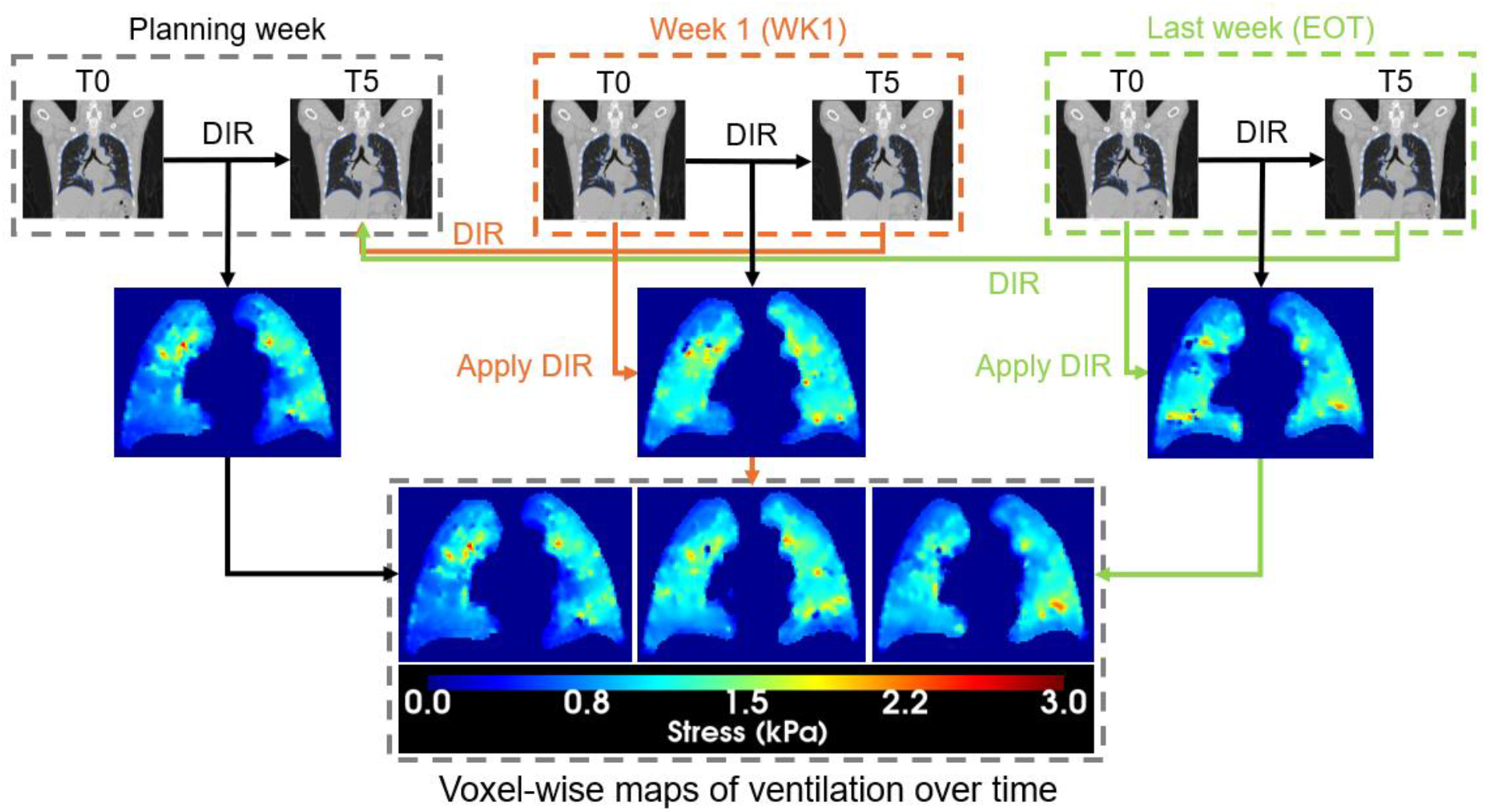
Diagram of workflow to generate voxel-wise ventilation maps at multiple timepoints. The three timepoints shown are planning (black), 1 week after treatment (orange), and the last week of treatment (green). Deformable image registrations (DIRs) are shown with arrows pointing to the reference exam. Each week has a 4-dimensional computed tomography (4DCT) scan, and the inhale (T0) and exhale (T5) phase scans were used to generate ventilation maps.

The internal gross tumor volume (iGTV), contoured on the average intensity scan of the planning 4DCT to account for tumor motion, was obtained from the clinical plan. All iGTV contours were expanded by 7mm to account for uncertainty in tumor boundary and potential for microscopic disease [45], and removed from the ventilation maps. Planned dose was extracted from the planning 4DCT T5 images. For proton patients, a clinically commissioned Monte Carlo algorithm [46] was employed to calculate planned physical dose using relative biological effectiveness (RBE) of 1.1.

### Image processing and data analysis

For each patient, voxel-wise ventilation change at WK1 and EOT was calculated, then binned according to planned dose in 5 Gy increments from 0 to 80 Gy. In each dose bin, median ventilation change was calculated for each patient. Patients were stratified based on modality and an increase or decrease in ventilation at WK1 in greater than 56% of voxels. For example, if a patient experienced an increase in ventilation in greater than 56% of voxels within the lung, they were classified into the ventilation increase group. This threshold was determined by generating ventilation maps at planning and WK1 using T9 as the moving image instead of T0 as a reproducibility assessment of the process to stratify patients as “increase” or “decrease”. For both the original ventilation maps with T0 and the ventilation maps with T9, the percentage of voxels that increased in ventilation was calculated. The median difference between percentages was calculated to be 6%, and therefore a threshold of 6% was used for stratification. The patients that did not meet this criterion, for example by having 52% of voxels with increased ventilation, were classified into a separate “stable” group. This led to 6 patient groups: photon patients with an increase in ventilation at WK1, photon patients with a decrease in ventilation at WK1, photon patients with stable ventilation at WK1, proton patients with an increase in ventilation at WK1, proton patients with a decrease in ventilation at WK1, and proton patients with stable ventilation at WK1. For each group, the median ventilation change in each dose bin was calculated across patients. This population-based approach was taken to account for high variability among patients and reduce noise. In each group, the number of patients that were stratified similarly at EOT in a majority of voxels was calculated and the number of patients that developed Grade 2+ pneumonitis was calculated. Fisher’s exact tests were performed to compare the incidence of pneumonitis between increase and decrease groups for each radiation modality. Mann-Whitney U tests were performed in each dose bin to compare the ventilation changes against zero and determine whether the change was significant (p<0.05).

GTV reduction at WK1 and EOT was calculated for each patient to correlate tumor changes with ventilation changes in univariate analysis and a linear regression model. Since the iGTV was contoured only on the planning 4DCT for clinical purposes, an in-house segmentation model was used to automatically contour the GTV on the T5 phase of the planning, WK1, and EOT 4DCT scans. The GTV was segmented on the planning 4DCT for comparison with WK1 and EOT instead of using the iGTV contour to ensure consistency between contours. All contours were visually inspected for accuracy. GTV reduction at WK1 and EOT was calculated for each patient.

Univariate analysis was performed to correlate median ventilation change at EOT throughout the entire lung with clinical metrics and other factors for each patient. A p-value below 0.05 was considered statistically significant. Variables in the model were median ventilation change in the whole lung at WK1 4DCT, age, sex, GTV size at planning, GTV location (right upper lobe, right middle lobe, right lower lobe, left upper lobe, or left lower lobe), GTV change at WK1 4DCT, total lung volume, modality (IMRT or PSPT), prescription dose, dose delivered at WK1 4DCT, smoking status, smoking history, and total years smoking.

A linear regression model was developed to predict median ventilation at EOT throughout the whole lung (*V*_*EOT*_) using 7 input features: median ventilation of the whole lung at planning (*V*_*WKO*_), median ventilation of the whole lung at WK1 (*V*_*WK1*_), GTV reduction at WK1 in cm^3^ (*ΔGTV = GTV*_*WK1*_−*GTV*_*WKO*_), functional lung volume (*FV =* 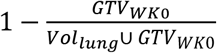where *Vol*represents the volume of the ipsilateral lung), a binary term to indicate whether the tumor was located in a lower or non-lower lobe (*L*where *L =* −1 for non-lower lobes and *L =* 1 for lower lobes), and two interaction terms (*ΔGTV* × *L*and *FV* × *L*). The binary term was included to assess the impact of tumor location near the airways by separating tumor location into lower (far from airways) and non-lower (near airways) lobes. The interaction terms were included to account for location-specific effects by allowing *ΔGTV* and *FV* to influence ventilation differently in lower lobes compared to other lobes. The model was fitted using an ordinary least squares model with an intercept using the following equation:

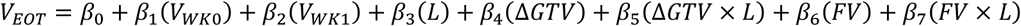

Model fit was summarized by R^2^ and by the correlation between predicted and observed median whole lung ventilation at EOT.

## Results

18 photon and 10 proton patients showed increased ventilation at WK1. For this cohort, increased ventilation at WK1 was predictive of increased ventilation at EOT for 13 (72%) and 8 (80%) of photon and proton patients respectively. 5 (28%) and 2 (20%) of photon and proton patients respectively developed Grade 2+ pneumonitis. 24 photon and 10 proton patients showed decreased ventilation at WK1, with 22 (92%) and 9 (90%) also decreasing at EOT respectively and 2 (8%) and 1 (10%) developing Grade 2+ pneumonitis respectively. Six photon and 3 proton patients were classified as having stable ventilation, with 3 (50%) and 2 (67%) patients also being classified similarly at EOT respectively. One out of 6 (17%) photon and 2 out of 3 (67%) proton patients developed Grade 2+ pneumonitis. For the increase and decrease groups, the ventilation changes at EOT were significantly different from zero in more dose bins for the photon patients (11/14 bins for increase, 14/14 bins for decrease) compared to proton patients (3/14 for increase, 5/14 for decrease). Ventilation changes at WK1 were significantly different from zero in all dose bins across increase and decrease groups as expected due to the stratification process. None of the dose bins for the stable group were significantly different from zero. Fisher’s exact tests comparing pneumonitis rates between increase and decrease groups returned insignificant p-values for photon (p=0.118) and proton (p=1.0) patients. The ventilation changes at each time point as a function of dose for each group are shown in Figure 3.

**Figure 3.**
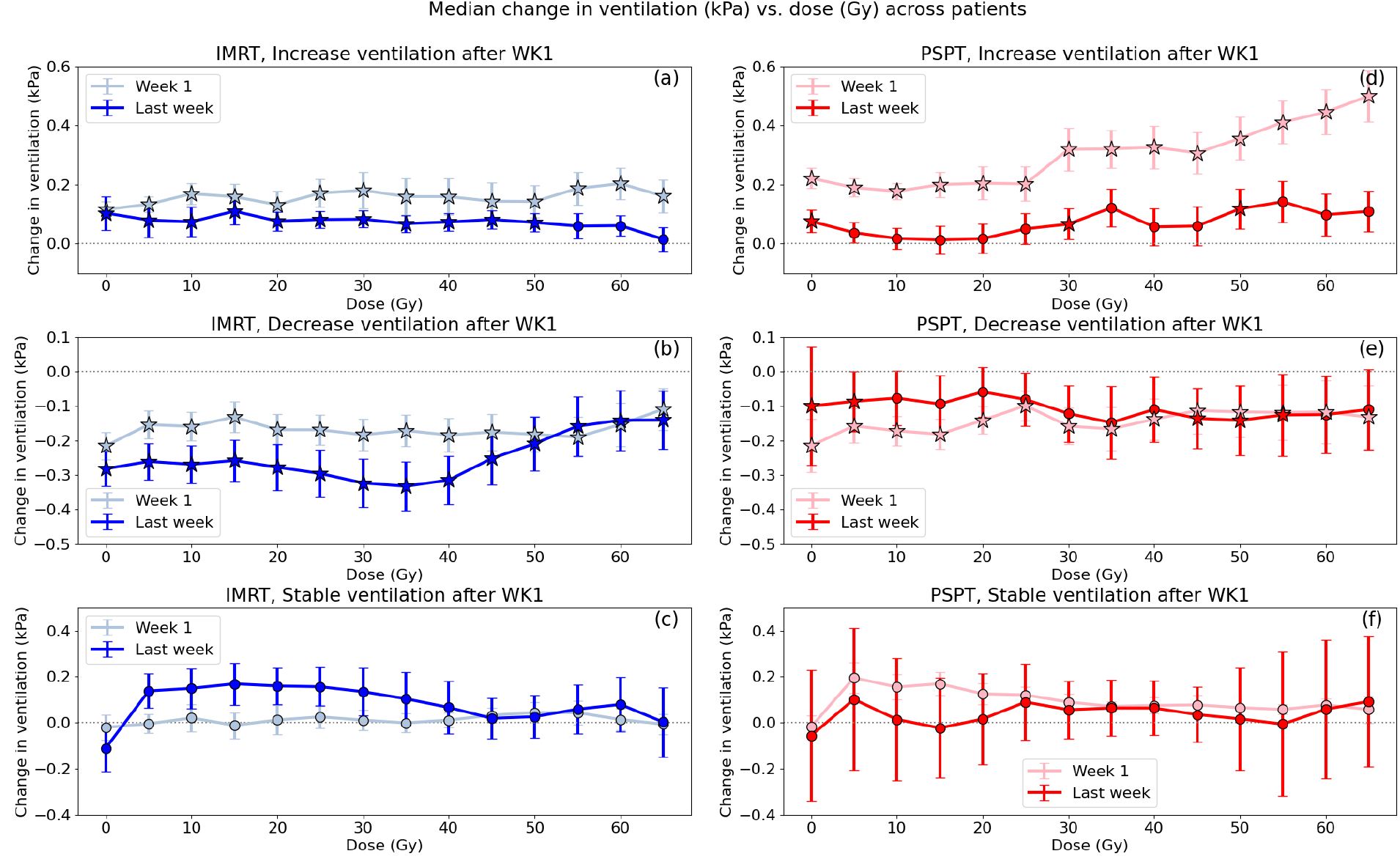
Plots of ventilation change after 1 week (light colors) and the last week (dark colors) of treatment for 6 patient groups. Ventilation change is shown on the y-axis, with positive values indicating an increase in ventilation. Planned dose bins from 0 to 70 Gy in 5 Gy increments are shown on the x-axis. Patients are split by photon (blue, figs. a-c) or proton (red, figs d-f) and increase (a, d), decrease (b, e), and stable (c, f) in ventilation at the first week of treatment (WK1). A positive value indicates that ventilation increased over time. Error bars represent standard error across patients. Stars indicate bins where Mann-Whitney U tests between the ventilation and zero values were significant (p<0.05).

The p-values for each independent variable from the univariate analysis are shown in Table 2, where bolded values indicate significance (p<0.05). Among all variables, only the median change in whole lung ventilation at the WK1 4DCT was significant (p=7.9E-9) in predicting median change in whole lung ventilation at EOT.

**Table 2.** Univariate analysis to predict Median Change in Whole Lung Ventilation at EOT.

|  |  |
| --- | --- |
| Independent Variable | P-value |
| Median change in whole lung ventilation at WK1 4DCT | **7.9 E -9** |
| GTV size at planning (cc) | 0.49 |
| Age (y) | 0.42 |
| Sex | 0.86 |
| Total lung volume (cc) | 0.91 |
| Modality (IMRT or PSPT) | 0.35 |
| GTV location | 0.68 |
| Smoking status (Y/N) | 0.83 |
| Smoking history (Y/N) | 0.26 |
| Total years smoking | 0.94 |
| Dose delivered @ WK1 (Gy) | 0.94 |
| Prescribed dose (Gy) | 0.44 |
| GTV reduction @ WK1 4DCT | 0.94 |
| Abbreviations: GTV = gross tumor volume, WK1 = week 1 of treatment, EOT = end of treatment, IMRT = intensity modulated radiation therapy, PSPT = passive scattering proton therapy |  |
| **Bold** indicates $p < 0.05$ | |

For the linear regression prediction model, the fitted coefficients were *β*_1_ *= 0*.*288*, *β*_2_ *=* 0.702, *β*_3_ *=* −0.874, *β*_4_ *=* 0.008, *β*_5_ *=* 0.009, *β*_6_ *=* 0.843, and *β*_7_ *=* 0.866, with intercept *β*_0_ *=* −0.884. The model achieved *R*^2^ *=* 0.65 which implies a Pearson correlation between predicted and observed *V*_*EOT*_ of √0.647 ≈ 0.804. Temporal predictors had the largest effect where *V*_*WK1*_ had the largest positive contribution with an additional independent contribution from *V*_*WKO*_. The marginal *ΔGTV* effect was *β*_4_ + *β*_5_ ≈ 0.018 per unit in lower lobes and *β*_4_−*β*_5_ ≈ −0.001 in non-lower lobes, and the marginal *FV* effect was *β*_6_ + *β*_7_ ≈ 1.709 per unit in lower lobes and *β*_6_−*β*_7_ ≈ −0.024 in non-lower lobes. A visual comparison of the observed vs. predicted *V*_*EOT*_ for each patient is shown in Figure 4.

**Figure 4.**
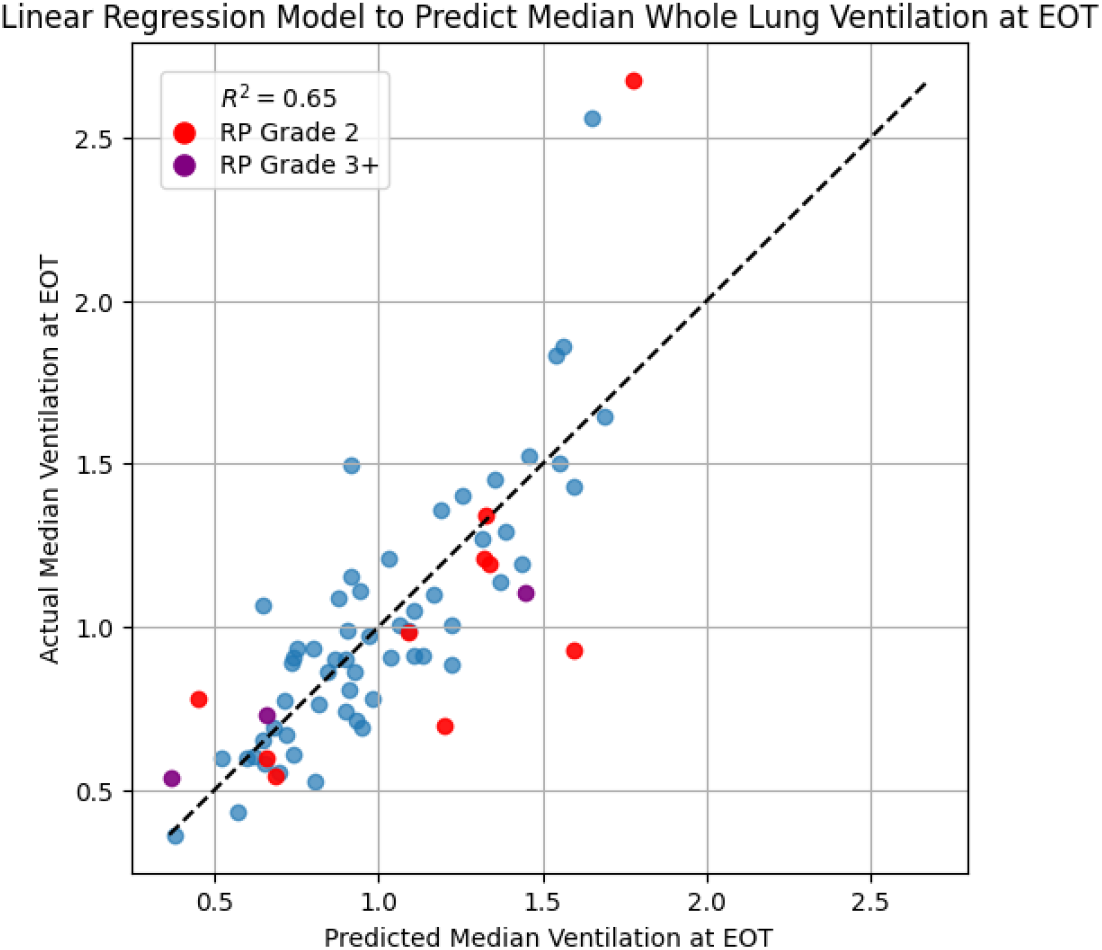
Linear regression model trained using 7 features to predict median whole lung ventilation at end of treatment (EOT). Predictions for each patient are shown on the x-axis and the actual ventilation at EOT is shown the y-axis, with the dashed line representing perfect prediction. Each marker represents 1 patient. Red markers indicate patients that developed Grade 2 pneumonitis (RP) and purple markers indicate patients that developed Grade 3 or higher RP.

## Discussion

In this study, changes in ventilation after WK1 were found to be predictive of changes at EOT through multiple analyses. Patients that were classified as “stable” during the stratification process at WK1 were less likely to be stratified similarly at EOT, indicating that the stratification process was less predictive for these patients. Univariate analysis in particular showed that only WK1 ventilation change can predict EOT change while no other clinical metrics can predict for it. For proton patients in particular, the ventilation change at EOT for both increase and decrease groups was not significantly different from zero in a majority of dose bins. While this could be due to a smaller number of proton patients compared to photon patients, it may also indicate that the higher normal tissue sparing from proton therapy results in less extreme impacts on ventilation.

The linear regression model indicated some dependency of the ventilation at EOT on the location of the tumor. The negative value for *β*_3_ indicates a lower baseline *V*_*EOT*_ for non-lower lobe tumors, suggesting that when tumors are located in lower lobes farther from the airways, the ventilation at EOT is higher. Interaction terms revealed a strong location-dependent modulation through the marginal effects for *ΔGTV* and *FV*, where the lower lobes had larger marginal effects compared to non-lower lobes. The predictions for two of the patients deviated substantially from the actual values for ventilation at EOT in Figure 4. For these patients, the ventilation at EOT were drastically different from ventilation at planning and WK1, which are the two most heavily weighted features in the model and may have led to the model’s inability to accurately predict the ventilation at EOT. The different ventilation at EOT may be due to the differences in ipsilateral lung volume between inhale and exhale, which were similar at planning and WK1 but doubled at EOT for both patients. These differences may be a result of an improvement in breathing ability due to tumor response, but further work is required to characterize the exact relationship between tumor location, lobe ventilation, and whole lung ventilation.

No dose dependency was observed for ventilation changes at WK1 and EOT. This is consistent with what was shown in Vinogradskiy et al. 2012 [36], which found that ventilation does not change as a function of dose. In our study, patients were found to both increase and decrease in ventilation, which is consistent with what was shown in Kipritidis et al. 2015 [35]. Our method of calculating ventilation was different from both of these studies, but similar results were still found. It is also worth noting that both of these studies looked at percentile ventilation and similar results were observed with the raw, non-normalized data from our study. While traditional CT ventilation imaging studies calculate percentile ventilation [12, 26, 27, 29, 47, 48] to normalize the data, we avoided normalizing by percentile in order to assess the raw, measured values of stress and interpret the results more clearly. However, it is important to note that patient breathing will likely vary over the course of treatment, especially when considering tumor response and reductions to airway obstruction, and further work with normalized data is needed.

Interestingly, patients that increased in ventilation at WK1 were more likely to develop pneumonitis, although the Fisher’s exact tests did not demonstrate significance when compared to the incidence rates for patients that decreased in ventilation at WK1. A previous study by Tucker et al. demonstrated that smokers are less likely to develop pneumonitis [49], potentially due to higher resistance to inflammation or decreased sensitivity to typical symptoms of pneumonitis, and a similar phenomenon may be in effect here with patients that decrease in ventilation at WK1. It is worth noting, however, that our study was limited by the small number of patients that developed pneumonitis, which can be visualized in Figure 4, and future studies should incorporate more patients.

An additional limitation of this study is the imbalance between the number of photon and proton patients. Due to the imaging data requirements for this study, a large percentage of the original cohort of proton patients had to be excluded, which limited a direct comparison between photon and proton cohorts. Future work will seek to include more proton patients with the necessary imaging data and further explore the clinical impact of the ventilation changes observed here with pneumonitis and patient-reported outcomes to demonstrate potential for adaptive radiotherapy.

## Conclusion

We have developed a metric that predicts overall change in ventilation at EOT using data at week 1 of treatment and found that no other clinical factors in our cohort demonstrated a correlation. This metric can identify patients that are likely to experience dramatic changes in their ventilation at EOT. Patients that increased in ventilation at WK1 were more likely to develop pneumonitis and future work will analyze the relationship between ventilation changes at EOT and pneumonitis more extensively. Analysis will also incorporate ventilation in separate lobes of the lung and in the GTV region.

## Data Availability

All data produced in the present study are available upon reasonable request to the authors.

## Acknowledgements

Research in this publication was supported in part by the National Institutes of Health/National Cancer Institute under award numbers P30CA016672, P01CA261669, and 1R01HL157273-01. Research was also supported by the Helen Black Image Guided Fund and resources of the Image Guided Cancer Therapy Research Program, the Tumor Measurement Initiative through the MD Anderson Strategic Initiative Development Program (STRIDE), the Quantitative Imaging Analysis Core, and the Summer Imaging Research Program under CATALYST at The University of Texas MD Anderson Cancer Center.

## Notes

### Competing Interest Statement

The authors have declared no competing interest.

### Author Declarations

IRB RCR03-0400 of The University of Texas MD Anderson Cancer Center gave ethical approval for this work

## References

1. Siegel, R.L., A.N. Giaquinto, and A. Jemal, Cancer statistics, 2024. CA Cancer J Clin, 2024. 74(1): p. 12–49.

2. Guo, Q., et al., Current treatments for non-small cell lung cancer. Front Oncol, 2022. 12: p. 945102.

3. Jang, J.Y., et al., Radiation pneumonitis in patients with non-small-cell lung cancer receiving chemoradiotherapy and an immune checkpoint inhibitor: a retrospective study. Radiat Oncol, 2021. 16(1): p. 231.

4. Aiad, M., et al., Comparison of Pneumonitis Rates and Severity in Patients With Lung Cancer Treated by Immunotherapy, Radiotherapy, and Immunoradiotherapy. Cureus, 2022. 14(6): p. e25665.

5. Suga, K., Technical and analytical advances in pulmonary ventilation SPECT with xenon-133 gas and Tc-99m-Technegas. (0914-7187 (Print)).

6. Vinogradskiy, Y., CT-based ventilation imaging in radiation oncology. BJR|Open, 2019. 1(1): p. 20180035.

7. Reinhardt, J.M., et al., Registration-based estimates of local lung tissue expansion compared to xenon CT measures of specific ventilation. Med Image Anal, 2008. 12(6): p. 752–63.

8. Guerrero, T., et al., Dynamic ventilation imaging from four-dimensional computed tomography. Physics in Medicine & Biology, 2006. 51(4): p. 777.

9. Yamamoto, T., et al., Four-dimensional computed tomography pulmonary ventilation images vary with deformable image registration algorithms and metrics. Medical Physics, 2011. 38(3): p. 1348–1358.

10. Castillo, E., et al., Robust CT ventilation from the integral formulation of the Jacobian. Medical Physics, 2019. 46(5): p. 2115–2125.

11. Castillo, E., Y. Vinogradskiy, and R. Castillo, Robust HU-based CT ventilation from an integrated mass conservation formulation. Medical Physics, 2019. 46(11): p. 5036–5046.

12. Castillo, R., et al., Ventilation from four-dimensional computed tomography: density versus Jacobian methods. Phys Med Biol, 2010. 55(16): p. 4661–85.

13. Kipritidis, J., et al., The VAMPIRE challenge: A multi-institutional validation study of CT ventilation imaging. Med Phys, 2019. 46(3): p. 1198–1217.

14. Hegi-Johnson, F., et al., Evaluating the accuracy of 4D-CT ventilation imaging: First comparison with Technegas SPECT ventilation. Med Phys, 2017. 44(8): p. 4045–4055.

15. Kipritidis, J., et al., Validating and improving CT ventilation imaging by correlating with ventilation 4D-PET/CT using 68Ga-labeled nanoparticles. Medical Physics, 2014. 41(1): p. 011910.

16. Yamamoto, T., et al., Pulmonary Ventilation Imaging Based on 4-Dimensional Computed Tomography: Comparison With Pulmonary Function Tests and SPECT Ventilation Images. International Journal of Radiation Oncology*Biology*Physics, 2014. 90(2): p. 414–422.

17. Lombardo, J., et al., Prospective Trial of Functional Lung Avoidance Radiation Therapy for Lung Cancer: Quality of Life Report. (1879-355X (Electronic)).

18. Nourzadeh, H., et al., Pneumonitis Prediction Modeling of a Prospective 4DCT-Ventilation Functional Avoidance Clinical Trial. International Journal of Radiation Oncology*Biology*Physics, 2022. 114(3, Supplement): p. S60–S61.

19. Vinogradskiy, Y., et al., Results of a Multi-Institutional Phase 2 Clinical Trial for 4DCT-Ventilation Functional Avoidance Thoracic Radiation Therapy. International Journal of Radiation Oncology*Biology*Physics, 2022. 112(4): p. 986–995.

20. Yamamoto, T., et al., Four-Dimensional Computed Tomography Ventilation Image-Guided Lung Functional Avoidance Radiation Therapy: A Single-Arm Prospective Pilot Clinical Trial. International Journal of Radiation Oncology*Biology*Physics, 2023. 115(5): p. 1144–1154.

21. Yaremko, B.P., et al., Functional Lung Avoidance for Individualized Radiation Therapy: Results of a Double-Masked, Randomized Controlled Trial. International Journal of Radiation Oncology*Biology*Physics, 2022. 113(5): p. 1072–1084.

22. Bucknell, N.A.-O., et al., Single-arm prospective interventional study assessing feasibility of using gallium-68 ventilation and perfusion PET/CT to avoid functional lung in patients with stage III non-small cell lung cancer. (2044-6055 (Electronic)).

23. Khalil, A.A., et al., Personal innovative approach in radiation therapy of lung cancer-functional lung avoidance SPECT-guided (ASPECT) radiation therapy: a study protocol for phase II randomised double-blind clinical trial. BMC Cancer, 2021. 21(1): p. 940.

24. Kida, S., et al., CT ventilation functional image-based IMRT treatment plans are comparable to SPECT ventilation functional image-based plans. Radiother Oncol, 2016. 118(3): p. 521–7.

25. Faught, A.M., et al., Evaluating the Toxicity Reduction With Computed Tomographic Ventilation Functional Avoidance Radiation Therapy. Int J Radiat Oncol Biol Phys, 2017. 99(2): p. 325–333.

26. Faught, A.M., et al., Evaluating Which Dose-Function Metrics Are Most Critical for Functional-Guided Radiation Therapy. International Journal of Radiation Oncology*Biology*Physics, 2017. 99(1): p. 202–209.

27. Flakus, M.J., et al., Metrics of dose to highly ventilated lung are predictive of radiation-induced pneumonitis in lung cancer patients. Radiother Oncol, 2023. 182: p. 109553.

28. Kanai, T., et al., Evaluation of functionally weighted dose-volume parameters for thoracic stereotactic ablative radiotherapy (SABR) using CT ventilation. Phys Med, 2018. 49: p. 47–51.

29. O’Reilly, S., et al., Dose to Highly Functional Ventilation Zones Improves Prediction of Radiation Pneumonitis for Proton and Photon Lung Cancer Radiation Therapy. Int J Radiat Oncol Biol Phys, 2020. 107(1): p. 79–87.

30. Rankine, L.J., et al., Quantifying Regional Radiation-Induced Lung Injury in Patients Using Hyperpolarized (129)Xe Gas Exchange Magnetic Resonance Imaging. Int J Radiat Oncol Biol Phys, 2024. 120(1): p. 216–228.

31. Baschnagel, A.M., et al., A Phase 2 Randomized Clinical Trial Evaluating 4-Dimensional Computed Tomography Ventilation-Based Functional Lung Avoidance Radiation Therapy for Non-Small Cell Lung Cancer. (1879-355X (Electronic)).

32. Ding, K., et al., 4DCT-based measurement of changes in pulmonary function following a course of radiation therapy. Medical Physics, 2010. 37(3): p. 1261–1272.

33. Patton, T.J., et al., Quantifying ventilation change due to radiation therapy using 4DCT Jacobian calculations. Med Phys, 2018. 45(10): p. 4483–4492.

34. Wallat, E.M., et al., Modeling the impact of out-of-phase ventilation on normal lung tissue response to radiation dose. (2473-4209 (Electronic)).

35. Kipritidis, J., et al., Measuring interfraction and intrafraction lung function changes during radiation therapy using four-dimensional cone beam CT ventilation imaging. (2473-4209 (Electronic)).

36. Vinogradskiy, Y.Y., et al., Use of weekly 4DCT-based ventilation maps to quantify changes in lung function for patients undergoing radiation therapy. Med Phys, 2012. 39(1): p. 289–98.

37. Yamamoto, T., et al., Changes in Regional Ventilation During Treatment and Dosimetric Advantages of CT Ventilation Image Guided Radiation Therapy for Locally Advanced Lung Cancer. International Journal of Radiation Oncology*Biology*Physics, 2018. 102(4): p. 1366–1373.

38. Liao, Z., et al., Bayesian Adaptive Randomization Trial of Passive Scattering Proton Therapy and Intensity-Modulated Photon Radiotherapy for Locally Advanced Non-Small-Cell Lung Cancer. J Clin Oncol, 2018. 36(18): p. 1813–1822.

39. Cazoulat, G., et al., Mapping lung ventilation through stress maps derived from biomechanical models of the lung. Medical Physics, 2021. 48(2): p. 715–723.

40. Cazoulat, G., et al., Biomechanical deformable image registration of longitudinal lung CT images using vessel information. Phys Med Biol, 2016. 61(13): p. 4826–39.

41. He, Y., et al., Quantifying the Effect of 4-Dimensional Computed Tomography–Based Deformable Dose Accumulation on Representing Radiation Damage for Patients with Locally Advanced Non-Small Cell Lung Cancer Treated with Standard-Fractionated Intensity-Modulated Radiation Therapy. International Journal of Radiation Oncology*Biology*Physics, 2024. 118(1): p. 231–241.

42. Brock, K.K., et al., Accuracy of finite element model-based multi-organ deformable image registration. Med Phys, 2005. 32(6): p. 1647–59.

43. Maas, S.A., et al., FEBio: finite elements for biomechanics. (1528-8951 (Electronic)).

44. Brock, K.K., et al., Use of image registration and fusion algorithms and techniques in radiotherapy: Report of the AAPM Radiation Therapy Committee Task Group No. 132. Medical Physics, 2017. 44(7): p. e43–e76.

45. Grills, I.S., et al., Clinicopathologic analysis of microscopic extension in lung adenocarcinoma: defining clinical target volume for radiotherapy. Int J Radiat Oncol Biol Phys, 2007. 69(2): p. 334–41.

46. Yepes, P., et al., Comparison of Monte Carlo and analytical dose computations for intensity modulated proton therapy. Phys Med Biol, 2018. 63(4): p. 045003.

47. Kida, S., et al., CT ventilation functional image-based IMRT treatment plans are comparable to SPECT ventilation functional image-based plans. (1879-0887 (Electronic)).

48. Vicente, E., et al., Functionally weighted airway sparing (FWAS): a functional avoidance method for preserving post-treatment ventilation in lung radiotherapy. Phys Med Biol, 2020. 65(16): p. 165010.

49. Jin, H., et al., Dose-volume thresholds and smoking status for the risk of treatment-related pneumonitis in inoperable non-small cell lung cancer treated with definitive radiotherapy. (1879-0887 (Electronic)).

